# Machine Learning for Dynamic and Short-term Prediction of Preeclampsia Using Routine Clinical and Laboratory Data

**DOI:** 10.1101/2025.09.29.25336926

**Authors:** Haoyang Li, Yaxin Li, Chengxi Zang, Weishen Pan, He S. Yang, Tracy B. Grossman, Zhen Zhao, Fei Wang

## Abstract

Preeclampsia (PE) is a leading cause of maternal and perinatal morbidity and mortality, yet its unpredictable onset and rapid progression hinder timely management. Existing prediction tools often rely on specialized biomarkers, static assessments, or limited study cohorts, impeding clinical utility and generalizability. We conducted a retrospective, multi-site cohort study including 58,839 pregnancies delivered at three NewYork-Presbyterian hospitals. Using routine information captured within the electronic health record (EHR), including blood pressure with other maternal characteristics, and routine laboratory tests, we developed extreme gradient boosting (XGBoost) based models to predict PE onset within 1-, 2-, and 4-week horizons across different gestational ages. Performance was assessed using nested cross-validation at the training site and externally validated through direct transfer, fine-tuning, and retraining strategies. Prediction accuracy increased from 28 to 34 weeks of gestational age, peaked at 34 weeks (AUC 0.863 at training; 0.808–0.834 at validation), declined at 38 weeks, and rebounded near delivery (AUC up to 0.890). Blood pressure was the most consistent predictor, while laboratory features such as albumin, alkaline phosphatase, and hematologic indices added value earlier, and demographic and obstetric factors gaining importance later. Dynamic short-term prediction of PE in late gestation is feasible using routine data. This pragmatic, scalable approach provides opportunities for early intervention and is adaptable across diverse healthcare settings.

## Introduction

Preeclampsia (PE) is a hypertensive disorder of pregnancy that affects 2-8% of pregnancies worldwide and remains a leading cause of maternal and perinatal morbidity and mortality ^1, 2, 3^. Beyond direct health consequences, PE drives significant healthcare utilization through preterm deliveries, neonatal intensive care admissions, and long-term follow-up for both mothers and infants ^4, 5, 6^. Its unpredictable onset and rapid progression pose critical challenges for obstetric care, making timely risk prediction a major unmet need.

A wide range of prediction strategies have been proposed, including maternal risk–based algorithms, biomarker panels, and machine-learning models ^7, 8, 9, 10^. Yet important limitations constrain their utility and implementation. Many approaches focus on PE prediction in the early pregnancy (the first trimester) to guide aspirin prophylaxis and enhanced surveillance, yet both predictive accuracy and preventive efficacy have shown only modest and inconsistent results across studies ^11, 12^. Others rely on specialized serum or ultrasound markers ^13, 14, 15, 16^, increasing cost and reducing accessibility in routine or resource-limited settings. Generalizability is also limited because most models were developed in a single cohort, even though the prevalence and severity of PE vary across racial and ethnic groups ^17, 18^, raising concerns of suboptimal performance when applied to different populations. Moreover, most existing tools assess risk at a specific time point and treat the risk static, rather than dynamically updating the risk as pregnancy progresses. Collectively, these limitations have hindered broad clinical adoption.

The timing of prediction is an underexplored dimension. Pregnancy is marked by dynamic physiologic changes, and PE can emerge abruptly, often in late gestation. While routine prenatal visits generate longitudinal clinical and laboratory data, most existing models predict only whether PE will occur at some stage of pregnancy, without estimating when onset is short-term ^8, 10, 19, 20^. Approaches explicitly designed around rolling observation and prediction windows can better capture short-term risk trajectories and provide actionable lead time for surveillance or intervention.

To address these gaps, we developed and validated Electronic Health Records (EHR)-based machine learning models for dynamic, short-term prediction of PE with the longitudinal EHR from over 58,000 pregnancies across three demographically distinct hospitals. Our model generates rolling risk estimates at different gestational age windows (28 to 40 weeks), forecasting PE onset within 1, 2, and 4 weeks. By adding a biomarker evaluation to routine clinical and laboratory assessments, our approach provides a practical, low-cost, and scalable method to assess pregnancies that may not be considered “high risk” for PE development. In summary, we show that PE in late gestation can be dynamically predicted with routinely available features, offering advanced actionable time for obstetric care.

## Results

### Cohort Characteristics

The study cohort was constructed from all pregnant individuals aged 18 years or older who delivered at three New York-Presbyterian hospitals: Weill Cornell Medical College (WCMC), Lower Manhattan Hospital (LMH), and Brooklyn Methodist Hospital (BMH). With the inclusion criteria described in the Method section, we obtained a cohort with a total of 58,839 pregnancies, 35,895 from WCMC, 8,664 from LMH, and 14,280 from BMH.

The cohort characteristics are summarized in Table 1, showing consistent patterns distinguishing PE pregnancies from healthy controls across all three hospitals. We observed that PE pregnancies were characterized by higher maternal age, greater proportions of Black race, nulliparity, multifetal gestation, and elevated body mass index (BMI) compared with controls. The distribution of PE patient counts across gestational weeks is presented in Supplementary Figure 1. Maternal age differences were statistically significant, with PE cases showing median ages approximately 1-2 years higher than healthy controls. For example, at WCMC, the median age was 35.0 years (Interquartile range, IQR 31.0–38.0) in PE cases versus 34.0 years (IQR 31.0–37.0) in controls (p <0.001), with similar patterns observed at LMH and BMH. Self-reported race distribution varied among three hospitals but all of them showed higher proportions of Black women among PE cases. The most pronounced differences were observed at WCMC (15.0% vs 6.5% Black women, p <0.001) and BMH (41.8% vs 21.8%, p <0.001). At LMH, Asian women comprised 33.3% of PE cases compared with 42.4% of healthy controls (p <0.001).

**Table 1.** Cohort Demographic and Baseline Characteristics.

| Variable | WCMC (N=35895) |  |  | LMH (N=8664) |  |  | BMH (N=14280) |  |  |
| --- | --- | --- | --- | --- | --- | --- | --- | --- | --- |
|  | PE (N=2227) | Control (N=3668) | p-value | PE (N=792) | Control (N=7872) | p-value | PE (N=1088) | Control (N=13192) | p-value |
| **Maternal Age at delivery(Median, IQR), year** | 35.0 (31.0 - 38.0) | 34.0 (31.0 - 37.0) | <0.001 | 35.0 (31.0 - 38.0) | 34.0 (32.0 - 37.0) | 2.94e-03 | 33.0 (28.0 - 36.0) | 31.0 (26.0 - 35.0) | <0.001 |
| **Race** |  |  |  |  |  |  |  |  |  |
| *White* | 1077 (48.4%) | 20424 (60.7%) | <0.001 | 254 (32.1%) | 2724 (34.6%) | 1.64e-01 | 408 (37.5%) | 7788 (59.0%) | <0.001 |
| *Black or African American* | 335 (15.0%) | 2178 (6.5%) | <0.001 | 117 (14.8%) | 566 (7.2%) | <0.001 | 455 (41.8%) | 2874 (21.8%) | <0.001 |
| *Asian* | 344 (15.4%) | 5659 (16.8%) | 1.01e-01 | 264 (33.3%) | 3336 (42.4%) | <0.001 | 34 (3.1%) | 559 (4.2%) | 9.13e-02 |
| *Other combinations* | 308 (13.8%) | 2917 (8.7%) | <0.001 | 100 (12.6%) | 704 (8.9%) | <0.001 | 131 (12.0%) | 1209 (9.2%) | 2.38e-03 |
| *Unknown* | 160 (7.2%) | 2467 (7.3%) | 8.35e-01 | 56 (7.1%) | 538 (6.8%) | 8.59e-01 | 58 (5.3%) | 755 (5.7%) | 6.39e-01 |
| **Smoker** |  |  |  |  |  |  |  |  |  |
| *Yes* | 242 (10.9%) | 2605 (7.7%) | <0.001 | 89 (11.2%) | 742 (9.4%) | 1.13e-01 | 64 (5.9%) | 597 (4.5%) | 4.86e-02 |
| *No* | 1972 (88.5%) | 30743 (91.3%) | <0.001 | 697 (88.0%) | 7081 (90.0%) | 9.65e-02 | 1016 (93.4%) | 12476 (94.6%) | 1.13e-01 |
| **Alcohol** |  |  |  |  |  |  |  |  |  |
| *Yes* | 20793 (61.8%) | 1399 (62.8%) | 3.29e-01 | 4657 (59.2%) | 498 (62.9%) | 4.61e-02 | 4119 (31.2%) | 377 (34.7%) | 2.11e-02 |
| *No* | 7562 (22.5%) | 543 (24.4%) | 3.80e-02 | 1968 (25.0%) | 178 (22.5%) | 1.27e-01 | 5070 (38.4%) | 411 (37.8%) | 6.92e-01 |
| **Multifetal gestation, n (%)** | 172 (7.7%) | 705 (2.1%) | <0.001 | 45 (5.7%) | 176 (2.2%) | <0.001 | 44 (4.0%) | 265 (2.0%) | <0.001 |
| **Delivery GA (Median, IQR), weeks** | 37.6 (36.6 - 39.1) | 39.3 (38.6 - 40.0) | <0.001 | 38.1 (37.1 - 39.4) | 39.3 (38.6 - 39.9) | <0.001 | 37.7 (36.7 - 39.3) | 39.6 (38.7 - 40.4) | <0.001 |
| **Cesarean delivery, n (%)** | 1231 (55.3%) | 10853 (32.2%) | <0.001 | 348 (43.9%) | 2275 (28.9%) | <0.001 | 551 (50.6%) | 3229 (24.5%) | <0.001 |
| **Baby weight (Median, IQR), g** | 2959.7 (2520.3 - 3365.1) | 3280.0 (2973.8- 3580.5) | <0.001 | 3050.4 (2656.3- 3424.6) | 3246.2 (2959.6- 3523.8) | <0.001 | 2979.5 (2489.0- 3418.9) | 3322.5 (2999.3- 3640.0) | <0.001 |
| **Nulliparity, n (%)** | 1524 (68.4%) | 18390 (54.6%) | <0.001 | 583 (73.6%) | 4439 (56.4%) | <0.001 | 600 (55.1%) | 5239 (39.7%) | <0.001 |
| **Pregavid BMI (Median, IQR), Kg/m<sup>2</sup>** | 28.6 (25.3 - 32.9) | 26.2 (23.9 - 29.4) | <0.001 | 28.3 (25.0 - 32.1) | 26.3 (24.0 - 29.5) | <0.001 | 32.6 (28.1 - 37.7) | 28.6 (25.4 - 32.9) | <0.001 |

*p value calculation:* p-values indicate group-wise comparisons between PE and controls within each site. For categorical variables, p-values were computed using a chi-squared test of independence based on contingency tables <sup>62</sup>. For continuous variables, comparisons were made using a two-sided Mann–Whitney U test <sup>63</sup>. p-values <0.001 are reported as "<0.001".

Besides, we observed that baseline obstetric risk factors were consistently elevated in PE cases. PE patients shown a distinct high-risk profile, with nulliparity significantly more prevalent (55-74% in PE cases versus 40-57% in healthy controls), multifetal gestation occurring two to three times more frequently, and pregravid BMI higher among PE cases across all hospitals. And pregnancy outcomes reflected the clinical burden of PE, characterized by earlier delivery (typically 1-2 weeks before term, with median gestational ages of 37.6-38.1 weeks in PE cases versus 39.3-39.6 weeks in healthy controls) and increased cesarean delivery rates (44-55% in PE cases compared to 24-32% in controls, all p <0.001).

### Prediction Performance Across Gestational Windows

The rolling window design revealed dynamic changes in prediction performance across different gestational windows, highlighting the need to divide the antepartum period into more granular time windows. We observed that prediction performance peaked around 34 weeks, declined at 38 weeks, and then rebounded at delivery. At the training site (WCMC, 2-week prediction window), Area under the Receiver Operating Characteristic Curve (AUC) rose from 0.803±0.061 at 28 weeks to 0.863±0.018 at 32 weeks and 0.866±0.015 at 34 weeks, before falling to 0.756±0.014 at 38 weeks and recovering to 0.850±0.020 at 40 weeks. Specificity at 90% sensitivity followed the same pattern, improving from 0.321±0.133 at 28 weeks to 0.569±0.127 at 34 weeks, dropping to 0.406±0.046 at 38 weeks, and rising again to 0.586±0.104 at 40 weeks. Positive predictive value (PPV) was low in early observation period (0.002±0.000 at 28 weeks) but increased to 0.018±0.002 at 34 weeks, peaked at 0.032±0.002 at 38 weeks.

The consistency of this performance pattern across external validation sites (LMH and BMH) demonstrated its generalizability. The external validation sites (LMH and BMH) exhibited similar trajectories. At the external validation sites (LMH and BMH), models were evaluated under different settings (see Method for details), including direct transfer application of the WCMC-trained model, local fine-tuning, and full retraining using site-specific data. At LMH (direct transfer, 2-week window), AUC improved from 0.775±0.099 at 28 weeks to peak performance 0.834±0.053 at 34 weeks, fell to 0.716±0.017 at 38 weeks, and rebounded strongly to 0.890±0.016 at 40 weeks. Similarly, at BMH, AUC improved from 0.749±0.092 at 28 weeks to 0.806±0.016 at 32 weeks and 0.808±0.014 at 34 weeks, dropped to 0.729±0.026 at 38 weeks, and rebounded to 0.808±0.029 at 40 weeks. And specificity demonstrated initial improvement from 28 to 34 weeks, followed by a decline at 38 weeks, and recovery at 40 weeks. PPV showed continuous improvement from 28 to 38 weeks, reaching its peak values at 38 weeks, before slightly decreasing at 40 weeks.

The largest performance gains occurred between 28 and 32–34 weeks, with average AUC improvements of 0.06, specificity increases of 0.10–0.25, and PPV gains of 0.006–0.015. Beyond 34 weeks, changes were less consistent: performance declined at 38 weeks but improved again at 40 weeks, highlighting a late gestation rebound. This rise–dip–rebound trajectory was consistent across the training and validation sites, indicating that predictive accuracy peaks around 34 weeks, diminishes before term, and recovers at delivery.

Extending the prediction horizon from 1 to 4 weeks produced smaller changes than shifting the observation window. For example, at WCMC with 𝑡_obs_=32 weeks, AUC was 0.855±0.033 for a 1-week prediction window, 0.863±0.018 for a 2-week prediction window, and 0.860±0.024 for a 4-week prediction window, while PPV increased from 0.005±0.001 to 0.026±0.004 as the prediction window lengthened. LMH and BMH showed similar stability, confirming that predictive performance was robust to forecast length.

When the WCMC-trained model was transferred directly to LMH and BMH, performance declined modestly, but local adaptation through fine-tuning or retraining restored most of the loss. At 𝑡_obs_=34 weeks (2 week prediction window), direct transfer performance at LMH was AUC 0.834±0.053, specificity 0.444±0.230, PPV 0.016±0.000, while fine-tuning improved these values to 0.868±0.054, 0.589±0.225, and 0.023±0.000, and retraining further to 0.871±0.017, 0.627±0.000, and 0.031±0.020. At BMH, AUC increased from 0.808±0.014 of direct transfer to 0.820±0.031 after retraining, with specificity improving from 0.438±0.000 to 0.456±0.128. Across GAs, direct transfer models at external sites lagged WCMC by 0.02–0.06 in AUC, but fine-tuning and retraining narrowed the gap to ≤0.01–0.02 and improved calibration, showing the importance of site-specific adaptation.

### Model Interpretability

Blood pressure was the dominant predictor across both early and late prediction windows ^21, 22, 23^. At 32–34 weeks, systolic blood pressure (SBP) and diastolic blood pressure (DBP) contributed the highest mean SHapley Additive exPlanations (SHAP) values, ranking first and second respectively. At 38–40 weeks, SBP and DBP again ranked first and second, remaining well above all other features. This consistency shows that blood pressure remained the most important predictor of PE regardless of gestational timing ^24, 25^.

At 32–34 weeks, laboratory features had greater influence compared to 38–40 weeks, supplementing blood pressure as key contributors. Albumin was the third-ranked feature, followed by alkaline phosphatase (ALP) and singleton pregnancy. Additional laboratory features such as globulin, Mean corpuscular hemoglobin concentration (MCHC), and mean corpuscular volume (MCV), as well as hematologic measures including hemoglobin and hematocrit, all ranked within the top 15. These results indicate that in the early third trimester, biochemical and hematologic markers provided substantial predictive value alongside blood pressure. While at 38–40 weeks, demographic and obstetric factors were becoming more important, while the importance of laboratory markers declined. Pregravid body mass index (BMI), number of term deliveries, and parity were all ranked in the top five, surpassing many laboratory features. White blood cell count (WBC) ranked 6th, maternal age ranked 7th, and gravidity ranked 10th. Meanwhile, albumin dropped from 3rd place at 32–34 weeks to 21st at 38–40 weeks, while creatinine improved from 24th to 17th place, and alanine transaminase (ALT) rose from 26th to 13th place. Overall, the relative weighting of feature groups shifted from laboratory-focused in early prediction to demographic-and obstetric-focused in late prediction, but with blood pressure consistently dominant. At 32–34 weeks, 5 of the top 10 features were laboratory or hematologic markers, versus only 2 at 38–40 weeks. Across both windows, blood pressure remained the leading predictor, but the supporting feature set transitioned from clinical laboratory values to maternal background characteristics as pregnancy progressed.

### Sensitivity Analyses

We conducted two sets of sensitivity analyses 1) if blood test features can significantly improve prediction performance; 2) if other machine learning models including logistic regression (LR) and random forest (RF) can perform better than our model. As shown in Figure 4, we observed that excluding blood test features and restricting models to blood pressure measurements along with static demographic, obstetric, and lifestyle variables resulted in performance drops, demonstrating that blood test features significantly enhanced prediction performance. We also observed that the other machine learning models LR and RF did not perform better than our XGBoost model, indicating that the prediction performance are not sensitive to the choice of machine learning models.

**Figure 1.**
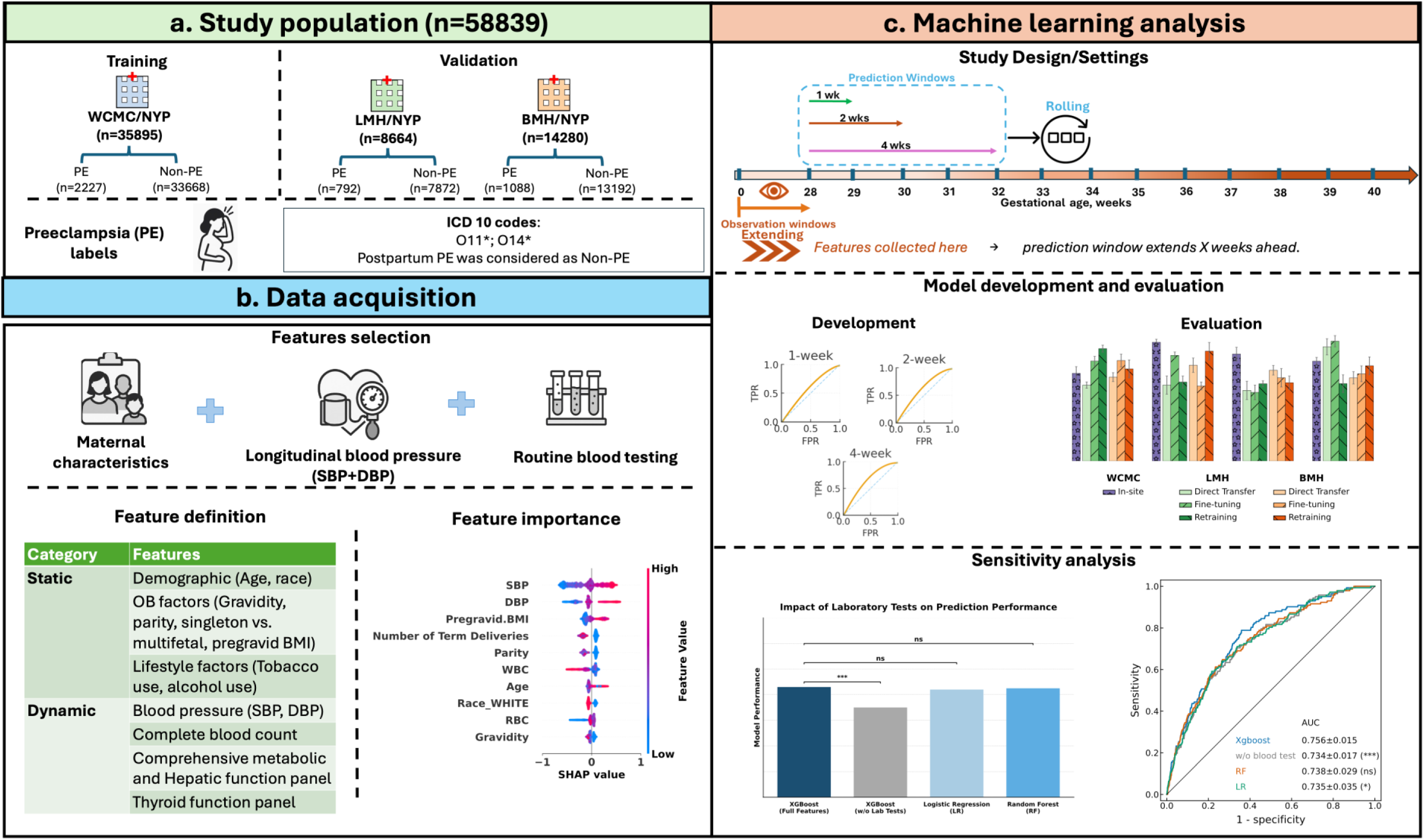
Study design and machine learning framework for preeclampsia (PE) prediction. (a) Cohorts included a training set from Weill Cornell (WC/NYP) and two external validation sets (LMH/NYP and BMH/NYP). (b) Input features comprised maternal characteristics, blood pressure, and routine laboratory panels; Feature importance were presented by SHAP value. (c) Models were developed across rolling observation windows (0–28 to 0–40 weeks) and short-term prediction horizons (1–4 weeks). XGBoost achieved the best performance compared with random forest and logistic regression, and sensitivity analyses showed that exclusion of laboratory data modestly reduced model accuracy.

**Figure 2.**
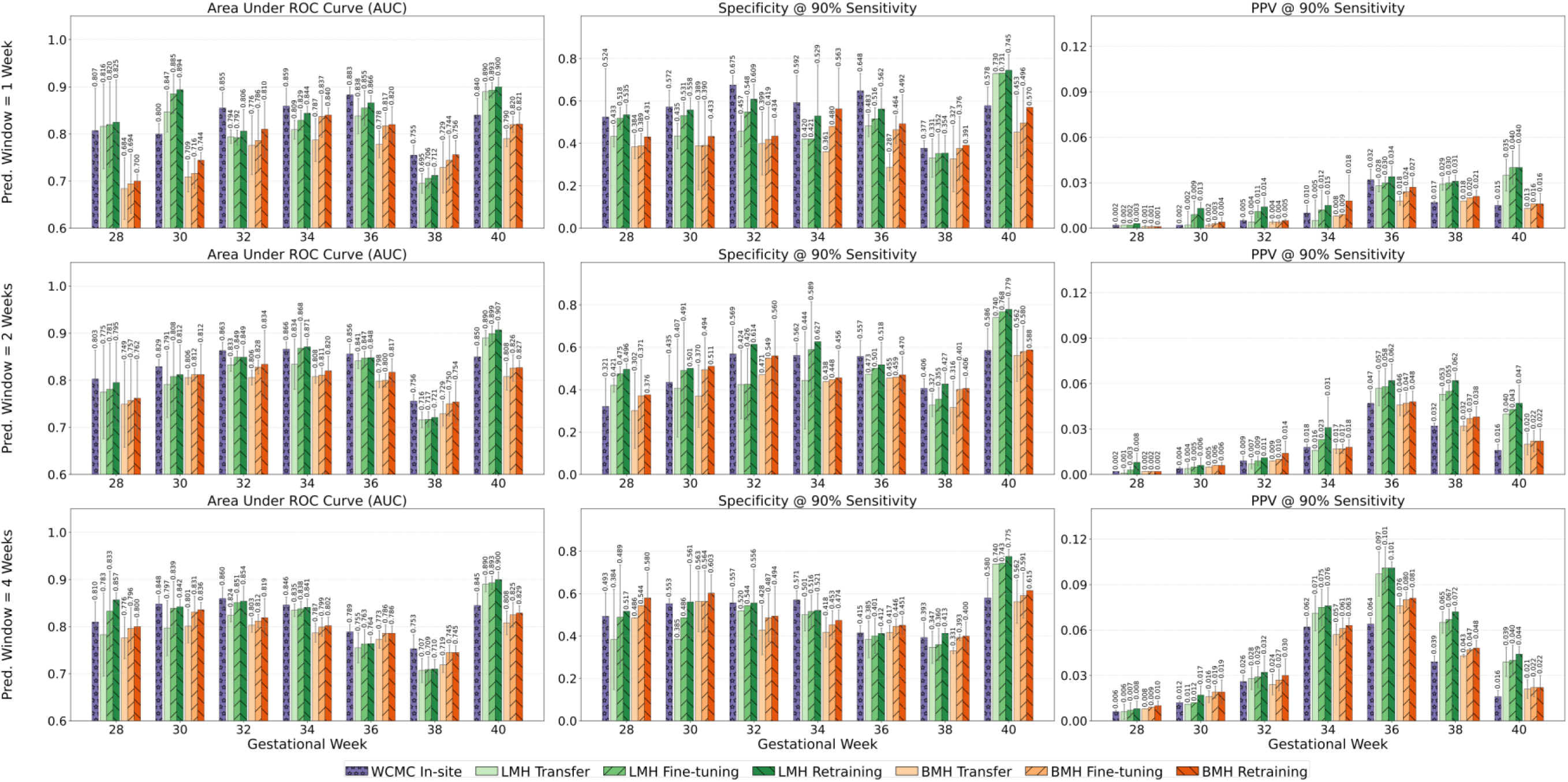
Prediction performance across gestational windows, prediction windows, and cross-site strategies. The prediction performance followed a consistent pattern across gestational windows, peaking around 34 weeks, declining at 38 weeks, then recovering at delivery across three different hospitals. Model performance was evaluated at index gestational weeks 28, 30, 32, 34, 36, and 38 with prediction windows of 1 week (top row), 2 weeks (middle row), and 4 weeks (bottom row). Each panel reports results from WCMC (in-site benchmark), LMH (external site), and BMH (external site) under three adaptation strategies: direct transfer, fine-tuning with local data, and complete model retraining. Metrics include AUC (left column), sensitivity at 90% specificity (middle column), and positive predictive value (right column). Across all sites, AUC and sensitivity improved as the observation window extended from 28 to 34 weeks and then plateaued through 38 weeks, while performance declined with longer prediction windows. Direct transfer resulted in lower external performance, whereas fine-tuning and retraining consistently restored accuracy toward in-site levels, demonstrating the value of local adaptation. AUC: Area Under the Receiver Operating Characteristic Curve; WCMC: Weill Cornell Medical College, LMH: Lower Manhattan Hospital, BMH: Brooklyn Methodist Hospital.

**Figure 3.**
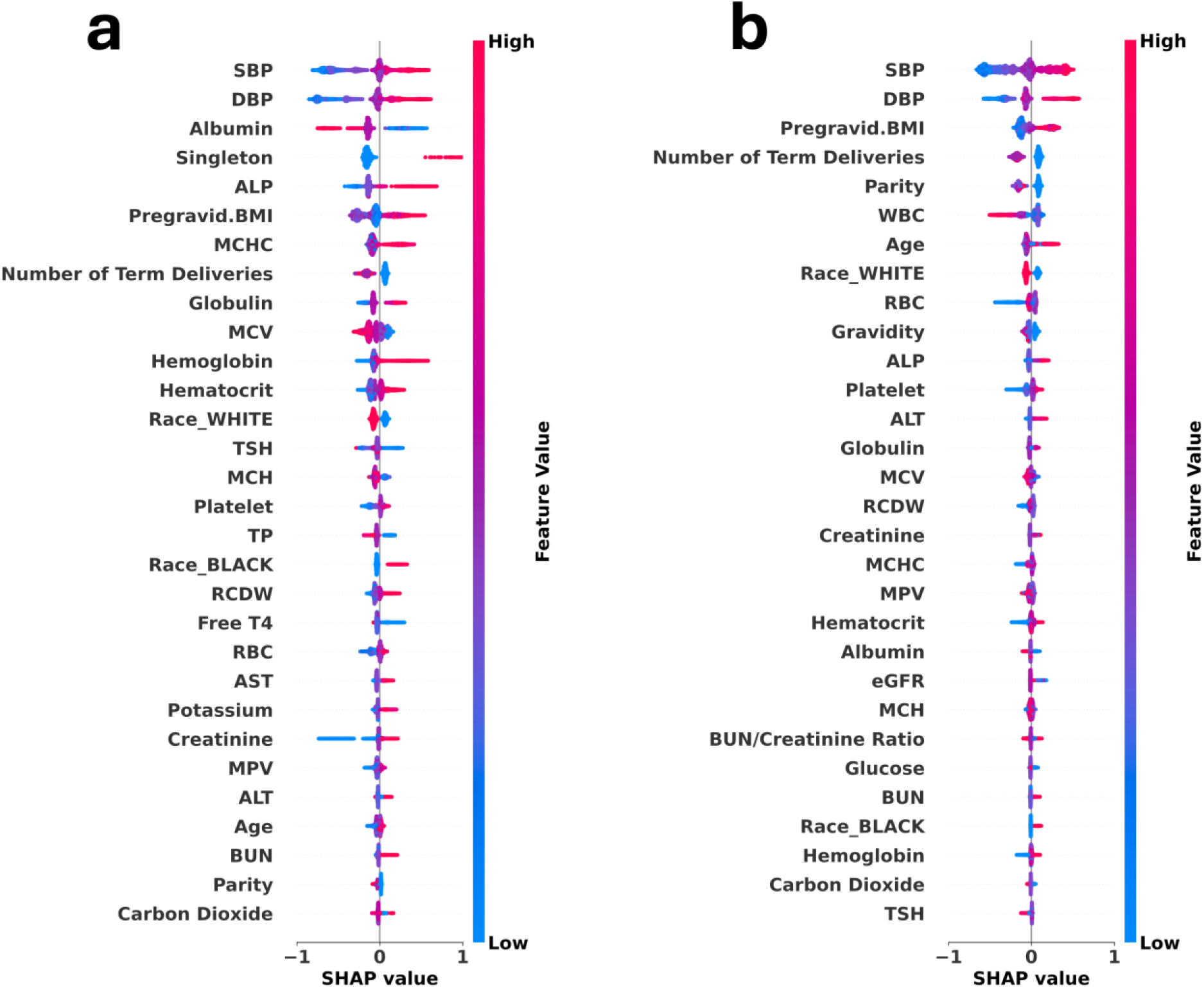
Top-30 global feature importance for PE prediction at early and late gestational stages. Global SHAP analyses were performed on XGBoost models trained at WCMC. (a) Observation window up to 32 weeks with a 2-week prediction window (32–34 weeks), representing early-onset prediction. (b) Observation window up to 38 weeks with a 2-week prediction window (38–40 weeks), representing late-onset prediction. Left panels show SHAP summary plots, where each dot represents an individual pregnancy and colors indicate feature values (red = high, blue = low). Right panels display the top 30 features ranked by mean absolute SHAP value. Across both settings, systolic and diastolic blood pressure, pregravid BMI, and obstetric history variables (parity, number of term deliveries) consistently ranked among the strongest predictors. Albumin and alkaline phosphatase contributed more prominently at 32 weeks, whereas age, white blood cell count, and gravidity gained importance by 38 weeks, highlighting stage-specific differences.

**Figure 4.**
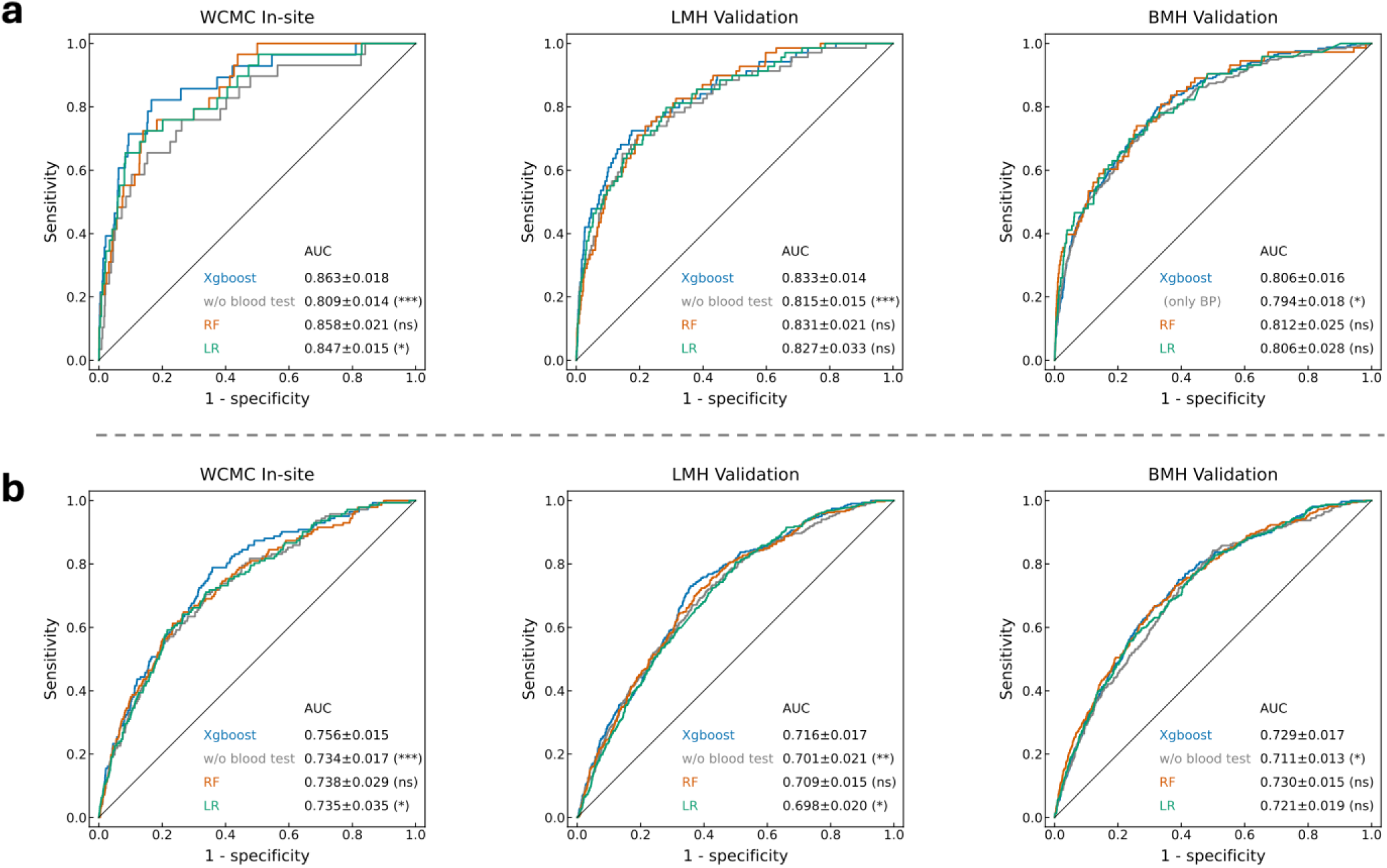
Sensitivity analyses of model variants across sites and gestational stages. Receiver operating characteristic (ROC) curves are shown for WCMC (in-site), LMH (external validation), and BMH (external validation). (a) Models trained with an observation window up to 32 weeks and a 2-week prediction window (32–34 weeks). (b) Models trained with an observation window up to 38 weeks and a 2-week prediction window (38–40 weeks). Default models (XGBoost, blue) were compared with models excluding blood test features (grey), logistic regression (orange), and random forest (green). Mean AUC ± standard error is reported in each panel. Statistical significance of performance differences was evaluated against the default XGBoost model using Welch’s t-test ^61^; results are annotated as ***: p < 0.001, **: p < 0.01, *: p < 0.05, and ns: not significant. Across all sites, exclusion of blood test features significantly reduced AUC, while logistic regression and random forest yielded performance comparable to XGBoost in most comparisons.

## Discussions

PE remains a critical obstetric challenge due to its unpredictable onset, rapid progression, and disproportionate burden in low-resource settings ^26, 27^. Most prior efforts have focused on first-trimester risk stratification to guide low-dose aspirin prophylaxis and enhanced monitoring ^28, 29^. While aspirin prophylaxis can reduce the incidence of preterm PE when initiated before 16 weeks at appropriate doses, its effect on late-onset and term PE, which are the majority of cases, has been minimal ^30, 31, 32^. As a result, current prediction strategies face a mismatch between clinical need and predictive capability. First-trimester prediction models achieve high performance for early-onset PE (AUC >0.8) but only modest accuracy for late-onset and term PE (AUC ∼0.6–0.7) Our study directly addresses these gaps by shifting the focus to late pregnancy, when most cases arise. By implementing rolling observation and prediction windows, our models not only identify who is at risk but also estimate when PE is likely to occur, within a 1–4-week horizon. This ability to capture both occurrence and timing provides clinically actionable lead time, aligning prediction with the timing of routine prenatal care and supporting timely intervention.

Presently, there is no feasible way to efficiently perform a longitudinal risk assessmentcontinually as the pregnancy progresses. A dynamic rolling-window approach addresses this need by continually updating the risk prediction using recent data, rather than treating risk as a fixed probability determined months in advance. Our rolling window design differs from existing studies, which typically treat PE risk as static, providing single estimates at fixed time points or across broad gestational stages ^8, 33^. While recent EHR-based studies have started to model pregnancy trajectories ^19, 34, 35^, they operate at coarse temporal resolution and focus on overall pregnancy outcomes rather than short-term risk estimation. Our framework generates risk estimation at a much finer temporal resolution in terms of GAs throughout late pregnancy, revealing a previously unrevealed pattern in predictive performance. We observed a consistent rise–peak–dip performance pattern across all sites, where AUC and specificity (at 90% sensitivity) improved from 28 to 34 weeks, declined at 38 weeks, then rebounded near delivery. This pattern aligns with prior works reporting that prediction of the predominant term PE remains challenging, with accuracy declining as the GA of onset advances ^36, 37^. Importantly, this rolling window design naturally stratifies patients by early-onset, preterm, and term PE without requiring predefined clinical labels, enhancing clinical applicability. Routine prenatal visits, which happen with more frequency as the pregnancy advances, generate a wealth of longitudinal data that can be used as part of a constantly-updating prediction model to give clinicians an updated estimated risk of the patient developing PE in the near future. This is clinically useful because it aligns with decision-making time frames: obstetric providers typically make week-by-week or visit-by-visit management decisions in late pregnancy.

The predictors we included in our model include maternal characteristics, blood pressure, and routine laboratory panels, which are widely available in routine EHR. This ensures practicality of the model in diverse clinical settings. Comparing the models built with different GAs and at different sites, blood pressure consistently emerged as the dominant predictor of PE, reflecting its central role in disease pathophysiology. Adding routine laboratory features can further enhance the model performance across sites and gestational windows. For example, at 32–34 weeks, inclusion of laboratory tests improved AUCs by ∼0.05 compared with models using blood pressure and other maternal characteristics alone, and similar gains were observed at later windows. Feature rankings also shifted with GA. At ∼32 weeks, protein-related and hematologic markers (albumin, globulin, ALP, and MCHC) were among the strongest contributors, consistent with prior reports linking early-onset PE to endothelial dysfunction and placental insufficiency ^38, 39, 40^. By 38 weeks, obstetric history, maternal age, and indicators of inflammation and hepatic stress (e.g., WBC, ALT) became more influential, aligning with the notion that late-onset PE is more strongly driven by maternal metabolic and inflammatory pathways ^40, 41^. Although the precise mechanisms remain incompletely understood, these shifts suggest that routine laboratory features not only enhance model performance but may also reflect biologically distinct processes across the spectrum of PE.

We have also tested the generalizability of our model. When the WCMC-trained model was directly applied to LMH and BMH, it maintained good discrimination performance (AUC>0.7 as defined in existing work ^42^), demonstrating the cross-site transferability of our model. We further evaluated local adaptation strategies to determine if they could enhance performance beyond direct transfer. Both strategies including model fine-tuning and model retraining provided performance gains, indicating that while the model generalizes well across sites, local adaptation can further optimize performance when site-specific data is available. Besides, the generalizability of our model was further demonstrated by the demographic diversity across hospitals. Each hospital served a distinct patient population. WCMC was majority White (∼60%), LMH had a larger proportion of Asian patients (∼40%), and BMH included the highest proportion of Black patients (∼23%). PE prevalence and severity are known to differ across racial and ethnic groups ^17, 43, 44^. Black women have consistently shown higher odds of PE and severity compared with White women ^45^, while Asian women often have lower relative risk ^17^. Applying the model across such demographic contrasts allowed us to evaluate its performance under real-world heterogeneity in both incidence and outcomes. Despite these differences, performance remained stable across all hospitals, demonstrating the generalizability of our model across diverse demographic groups.

This study has several limitations. First, it was retrospective and conducted within a single healthcare system, though across three demographically distinct hospitals. Validation in more diverse patient populations, particularly in low-and middle-income countries, will be essential. Second, although we analyzed more than 58,000 pregnancies, the longitudinal data had missing values and irregular sampling. While this reflects real-world practice, the large sample size helps mitigate these gaps, and our models were designed to rely on either median blood pressure or the most recent laboratory results together with static demographic and obstetric variables. Third, PE diagnoses were based on EHR documentation and ICD codes, which may introduce misdiagnosis, though the risk is partly offset by the large cohort size and the consistency across sites.

In summary, this study demonstrates that dynamic, short-term prediction of PE in late gestation is feasible using only routinely available clinical features. By providing advanced actionable time and relying on a simple, uniform feature set, our framework offers a pragmatic foundation for scalable implementation. These strengths highlight its potential to support more timely and accessible obstetric care across varied healthcare settings.

## Method

### Data and cohort

This study was approved by the Institutional Review Board of Weill Cornell Medicine (IRB#24-05027450). All analyses were performed on de-identified EHR data. The study was approved by the Weill Cornell Medicine Institutional Review Board with a waiver of informed consent for secondary use of de-identified EHR data.

We conducted a retrospective, multi-site cohort study of pregnant individuals who received prenatal care and delivered at three New York Presbyterian hospitals: WCMC, LMH, and BMH. The dataset includes structured clinical information spanning demographics, vital signs, laboratory results, and diagnosis codes, recorded as part of routine obstetric care. The study cohort was constructed by identifying pregnancies with a documented delivery date between October 2020 and May 2025. Inclusion criteria required patients to be 18 years or older with documented delivery records that included both delivery time and gestational age at delivery during the study period. After excluding 247 pregnancies (142 from WCMC, 22 from LMH, and 83 from BMH) due to missing delivery time or gestational age at delivery, the final cohort comprised 58,839 pregnancies: 35,895 from WCMC, 8,664 from LMH, and 14,280 from BMH. Each pregnancy was treated as a distinct observation. Multiple gestations were included, with no restrictions on parity. GA across the pregnancy was calculated based on the delivery date and delivery GA recorded in the EHR. Clinical variables were aligned to gestational week using date of service and delivery date/GA-derived reference timepoints. Pregnancies were identified by a clinician-documented delivery encounter; pregnancy onset (GA = 0 weeks, LMP-equivalent) was back-calculated as delivery date minus GA at delivery, and delivery GA/date/time were abstracted from the delivery record. PE was defined using ICD-10 diagnostic codes corresponding to antepartum PE, including both mild and severe subtypes (O11.x and O14.x). The time of PE onset was operationalized as the admission date of the first diagnosis code within the pregnancy episode. Vital signs and laboratory tests were retrieved from structured flowsheets and laboratory panels, respectively. Data available for model development included demographics (maternal age, race), obstetric characteristics (parity, gravidity, pregravid BMI, singleton vs. multifetal gestation), vital signs (blood pressure), lifestyle (tobacco or alcohol use) and laboratory results including complete blood count (CBC) panel, comprehensive metabolic panel (CMP), Hepatic and Thyroid functions. All features were time-stamped.

### Feature extraction and preprocessing

We summarized features used for model development in Supplementary Table 1. Demographic and obstetric characteristics, including maternal age, race, gravidity, parity, pregravid BMI, and pregnancy type (singleton vs. multifetal), were treated as fixed covariates, with lifestyle factors (tobacco/alcohol use) also incorporated. Blood pressure features were summarized as the median of all prenatal systolic and diastolic measurements within the observation window (defined in the Experimental Setting), minimizing the impact of outliers while reflecting overall maternal hemodynamics. For laboratory analytes, the most recent value within the observation window was used to capture the latest clinical status. Categorical variables were one-hot encoded, while continuous features were imputed with the median and categorical features with the mode, estimated within each training fold to prevent leakage. Tree-based models leveraged their native handling of missing values, whereas logistic regression used the imputed features directly. Continuous variables were z-standardized using statistics from the training split, with the same transformation applied to validation and test sets ^46^. Finally, the modeling features comprised the most recent laboratory results, median systolic and diastolic blood pressure, and static demographic, obstetric, and lifestyle variables including tobacco use and alcohol use.

### Experimental setting

We formulated the task as a binary classification problem anchored to GA. For each pregnant individual, we defined two key temporal intervals: (1) Observation window: the period from conception up to a cutoff week 𝑡_obs_, during which clinical and laboratory data are collected as inputs. (2) Prediction window: a future time interval of length Δ weeks, spanning [𝑡_obs_, 𝑡_obs_ + Δ], during which the model aims to forecast whether PE will occur. Let 𝑇 denote GA in weeks. The objective is to predict, at time 𝑡_obs_, whether a patient will develop PE during the subsequent prediction window. Patients who develop PE in [𝑡_obs_, 𝑡_obs_ + Δ] are labeled as positive; those who do not develop PE by the end of the prediction window are labeled as negative. Formally, let 𝐱^(𝒕obs)^ represent the feature vector for the individual 𝑖, constructed from all available data up to 𝑡_obs_. The outcome variable is defined as:

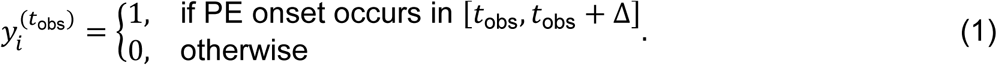

We then trained a probabilistic binary classifier 𝑓 to estimate the risk score for each patient:

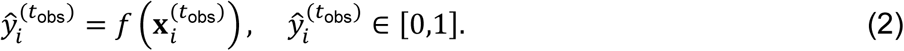

### Model development and validation

We implemented extreme gradient boosting (XGBoost) as the primary modeling framework ^47^. All experiments followed a nested cross-validation design, with both the outer and inner loops based on stratified 5-fold splits ^48^. The outer loop consisted of 5-fold cross-validation, where the dataset was split into five patient-level folds. In each round, one fold was held out for evaluation, and the remaining four folds were used for training and hyperparameter tuning. The inner loop used a 5-fold split on the training data to identify optimal hyperparameters via grid search on the validation set. The final reported metrics were averaged across the five outer test folds. All preprocessing steps, including imputation, feature scaling, and model training, were performed within each training fold to avoid information leakage ^49^. The chosen hyperparameters from validation were then applied to retrain the model on the full training data before evaluation on the outer test fold. Because the incidence of PE was relatively low, we applied sample reweighting to balance positive and negative classes with the class-weighted loss functions in XGBoost, where the positive class weight was set proportional to the inverse prevalence ^50^:

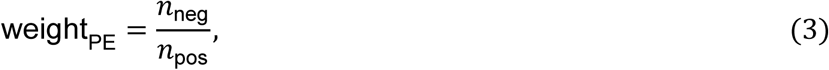

with 𝑛_pos_ and 𝑛_neg_ denoting the number of positive and negative samples in training folds, respectively. In sensitivity analyses, we compared other machine-learning prediction models, including regularized logistic regression and random forests, ^51, 52^ under the same experimental setup as the primary analysis, enabling direct comparisons.

### Cross-site evaluation strategies

To evaluate generalizability across healthcare systems, we designed four complementary strategies for model transfer and adaptation ^53, 54, 55^. (1) **In-site testing**: Models were trained and validated within the same hospital using nested cross-validation; this was applied only at WCMC, which served as the internal performance benchmark. (2) **Direct transfer** ^56^: Models trained at WCMC, where the largest number of pregnancies was available, were directly applied to LMH and BMH without modification. This represented the simplest form of external validation but was expected to be most sensitive to distributional differences in demographics, laboratory availability, and clinical practice patterns. (3) **Fine-tuning**: The model architecture and hyperparameters tuned at WCMC were fixed, with additional trees added to the existing XGBoost model using site-specific training data from LMH or BMH. This preserved the source model’s structure while adapting to local feature distributions, requiring moderate computational and data resources. (4) **Retraining**: Complete model redevelopment was performed separately at LMH and BMH, including new hyperparameter optimization within each site. This maximized alignment to local data but required the greatest resources and reduced cross-site comparability. Performance under these strategies was assessed for each observation window and prediction window. Overall, in-site testing reflected best-case accuracy within a given site, direct transfer represented external portability without adaptation, and fine-tuning or retraining quantified the benefits of local adaptation.

### Feature importance analysis

To interpret model predictions, we applied SHAP to the trained XGBoost models ^57^. SHAP values were calculated for each feature in the outer test folds of the nested cross-validation pipeline, ensuring that attribution was based only on held-out data. Global feature importance was summarized using the mean absolute SHAP value across all individuals. We examined feature contributions separately for early-onset prediction and late-onset prediction. Early-onset prediction was represented by an observation window up to 32 weeks with a 2-week prediction window (32–34 weeks), while late-onset prediction was represented by an observation window up to 38 weeks with a 2-week prediction window (38–40 weeks). This design allowed us to assess whether the importance of specific features shifted between earlier and later stages of pregnancy. The feature set included demographics (e.g., age, race/ethnicity), obstetric history (e.g., gravidity, parity, pregravid BMI, pregnancy type), lifestyle factors (tobacco and alcohol use), vital signs (median SBP and DBP), and laboratory tests (CBC panel, CMP, Hepatic and Thyroid functions), see Supplemental TableS1. For each model, the ranked feature list highlighted both clinical covariates and laboratory markers that contributed most strongly to predictions of PE risk. Results from SHAP analyses were visualized to compare the global feature rankings between early and late prediction tasks. These comparisons enabled identification of features with consistent predictive value across gestation, as well as features whose contributions were stage-specific.

### Evaluation metrics

Model performance was quantified using three complementary metrics: (1) Area under the receiver operating characteristic curve (AUC) ^58^. AUC measured overall discrimination between pregnancies that did and did not develop PE within the prediction window. This metric is independent of prevalence and provides a global assessment of classification performance. (2) Specificity at 90% sensitivity ^55, 59^: To reflect the clinical priority of minimizing missed cases, we fixed the operating point at 90% sensitivity and reported the corresponding specificity, capturing the ability to detect true PE cases while limiting false positives to 10% of unaffected pregnancies. Thresholds were determined within the inner cross-validation loop and applied to the corresponding outer test fold. (3) Positive predictive value (PPV) at 90% sensitivity ^60^: PPV measured the proportion of true PE cases among pregnancies classified as high risk. Because PE prevalence is relatively low, PPV served as a critical indicator of clinical utility beyond overall discrimination.

Performance was summarized at each gestational index week and for each prediction window (1, 2, and 4 weeks). These metrics were reported consistently across in-site testing, direct transfer, fine-tuning, and retraining strategies, enabling direct comparison of discrimination, detection capability under fixed specificity, and precision across sites.

### Sensitivity analyses

To examine the robustness of our findings, we performed a series of sensitivity analyses. First, we evaluated the impact of excluding laboratory tests from the feature set. In this setting, models were trained using only demographic variables, obstetric history, and median blood pressure. This analysis tested whether routinely collected non-laboratory data were sufficient for risk prediction and quantified the incremental value of blood test features. Second, we compared XGBoost with two baseline classifiers: logistic regression and random forest. Both models were trained and validated on the same cross-validation folds and feature sets as XGBoost. Performance under these sensitivity settings was assessed using the same evaluation metrics.

## Data Availability

This study analyzed de-identified electronic health record data from three hospitals within the NewYork-Presbyterian health system. Owing to patient privacy and institutional regulations, these data are not publicly available now. Access might be provided to qualified researchers upon reasonable request to the corresponding author and subject to approval by the Weill Cornell Medicine Institutional Review Board.

## Code Availability

The primary repository is hosted on https://github.com/lihy96/PreeclampsiaPrediction. The experiments were conducted using Python 3.10, using open-source libraries, including scikit-learn (version 1.5.1), XGBoost (version 2.1.1), and SHAP (version 0.48.0). All implementation details, including preprocessing scripts, model training, and hyperparameter configurations, are documented within the repository.

## Acknowledgements

The study was not supported by any external funding.

## Author Contributions

F.W. and Z.Z. conceived the initial idea. H.L. conceived the method and designed the algorithmic techniques. Y.L. contributed to data curation and methodology. H.L. wrote the codes and performed the computational analysis. H.L. and Y.L. drafted the initial manuscript, with critical revisions by F.W., Z.Z, and C.Z. W.P., H.Y., and T.G. reviewed the manuscript and provided suggestions. F.W. and Z.Z. supervised the project. All authors reviewed, provided feedback, and approved the final manuscript.

## Supplementary Materials

## Supplementary Table

**Supplementary Table 1.** Summary of features used for model development.

| Category | Sub-Category | Features |
| --- | --- | --- |
| Clinical characteristics | Demographics | Age; Race |
|  | Obstetric factors | Gravidity; Parity; Number of term deliveries; Pregravid BMI; Singleton pregnancy |
|  | Lifestyle | Tobacco use; Alcohol use |
|  | Blood pressure | Systolic blood pressure (SBP); Diastolic blood pressure (DBP) |
| Laboratory tests<br>(contains calculated results) | Complete blood Count panel | WBC; RBC; Hemoglobin; Hematocrit; Platelet count; MCV; MCH; MCHC; RDW; MPV; NRBC |
|  | Comprehensive metabolic and Hepatic function panels | Calcium; Sodium; Potassium; Chloride; Bicarbonate; Anion gap; Glucose; Creatinine; BUN; BUN/Creatinine ratio; eGFR; Albumin, Total protein; ALT; AST; ALP; Total bilirubin; Direct bilirubin; Indirect bilirubin; Globulin; Albumin/Globulin ratio |
|  | Thyroid function | TSH; Free T4 |

## Supplementary Figure

**Supplementary Figure 1.**
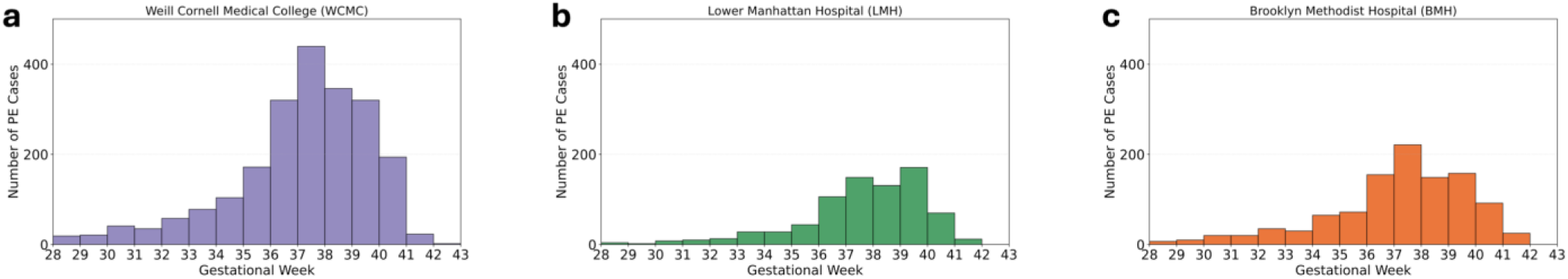
**The distribution of PE patient counts across gestational weeks of three hospitals.**

**Supplementary Figure 2.**
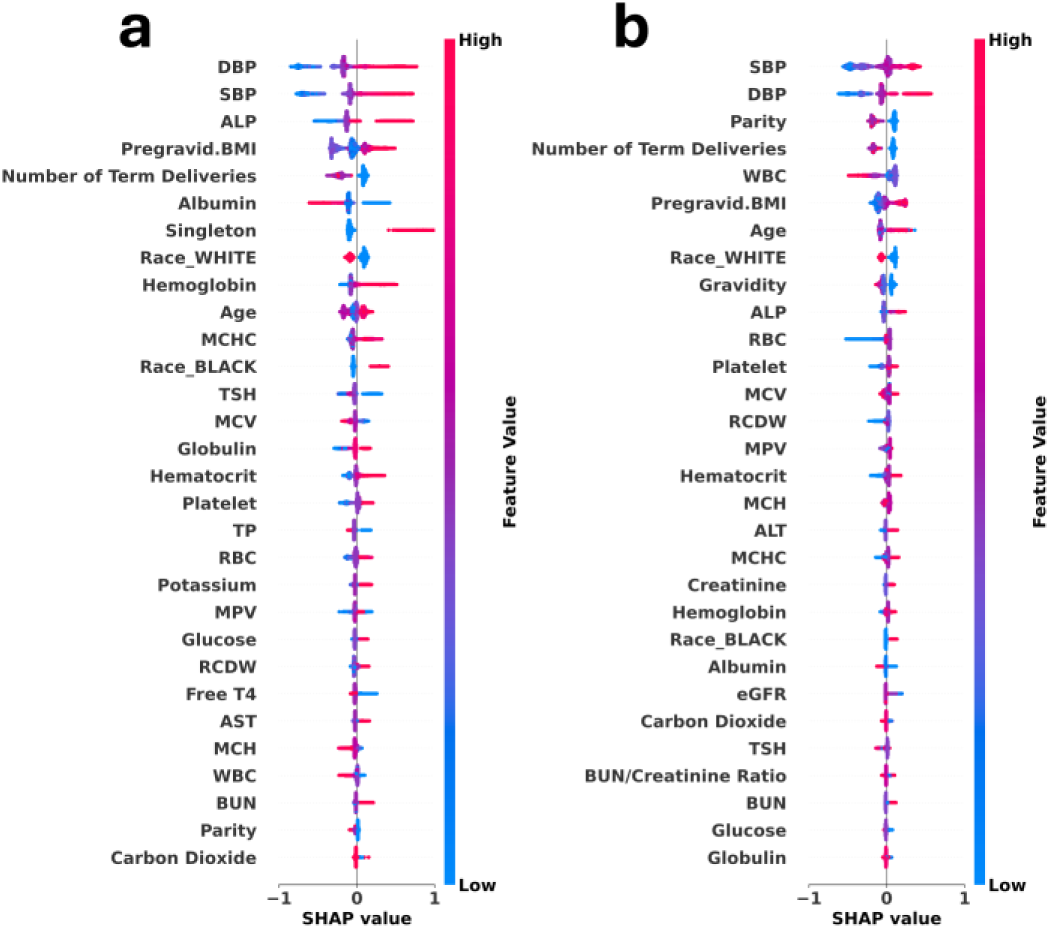
Top-30 global feature importance for PE prediction at early and late gestational stages. Global SHAP analyses were performed on XGBoost models (LMH direct transfer). (a) Observation window up to 32 weeks with a 2-week prediction window (32–34 weeks), representing early-onset prediction. (b) Observation window up to 38 weeks with a 2-week prediction window (38–40 weeks), representing late-onset prediction. In these SHAP summary plots, each dot represents an individual pregnancy and colors indicate feature values (red = high, blue = low).

**Supplementary Figure 3.**
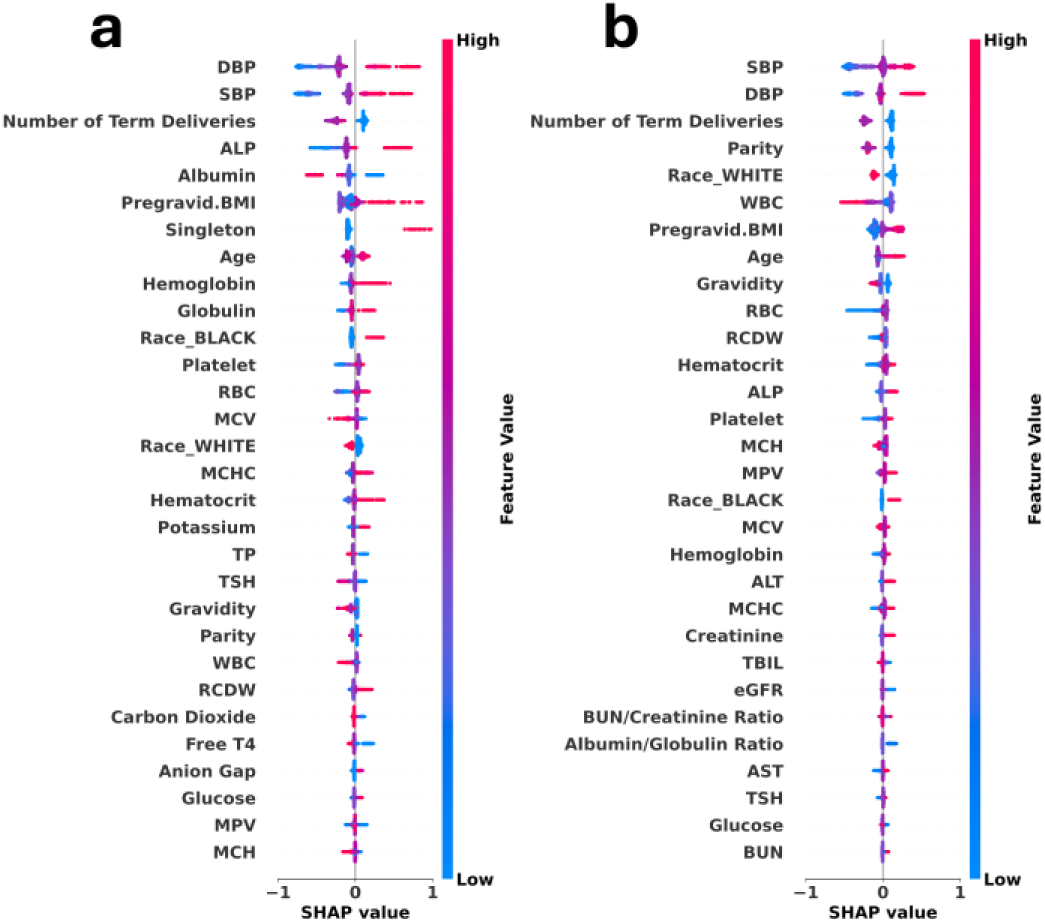
Top-30 global feature importance for PE prediction at early and late gestational stages. Global SHAP analyses were performed on XGBoost models (LMH fine-tuning). (a) Observation window up to 32 weeks with a 2-week prediction window (32–34 weeks), representing early-onset prediction. (b) Observation window up to 38 weeks with a 2-week prediction window (38–40 weeks), representing late-onset prediction. In these SHAP summary plots, each dot represents an individual pregnancy and colors indicate feature values (red = high, blue = low).

**Supplementary Figure 4.**
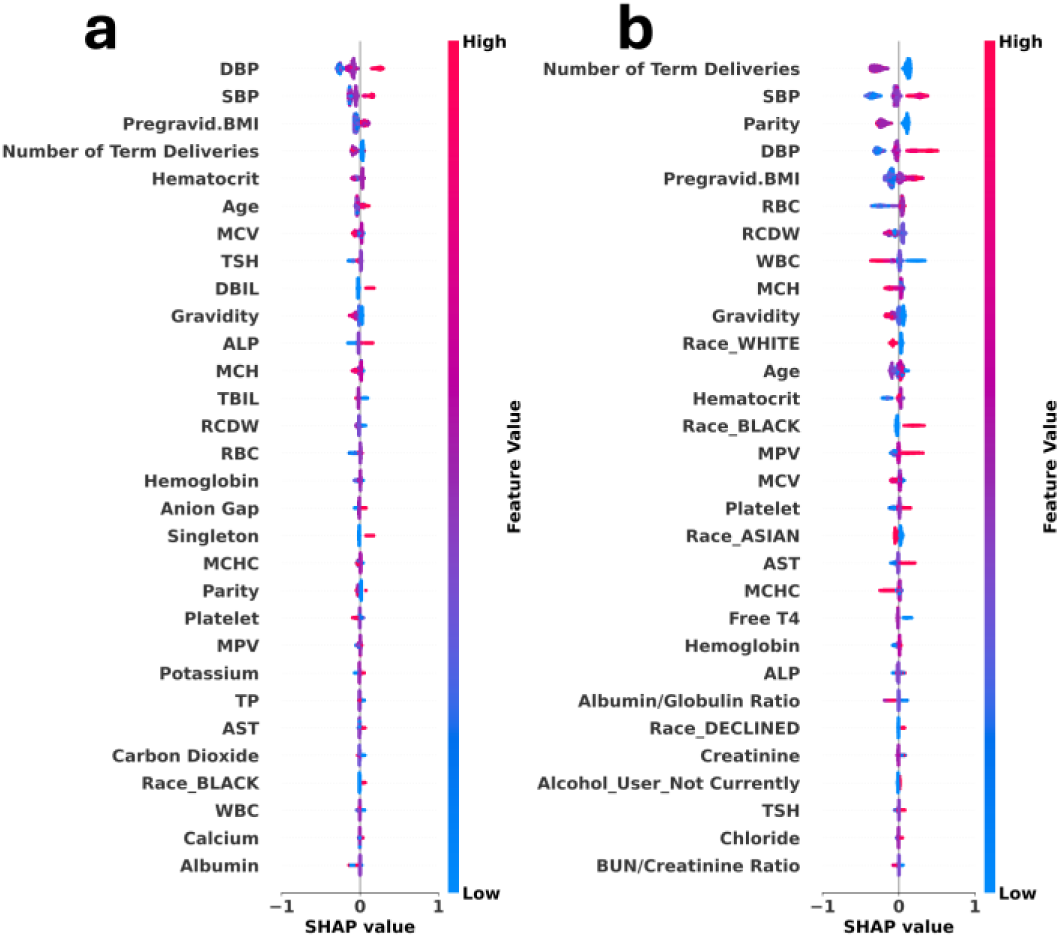
Top-30 global feature importance for PE prediction at early and late gestational stages. Global SHAP analyses were performed on XGBoost models (LMH retraining). (a) Observation window up to 32 weeks with a 2-week prediction window (32–34 weeks), representing early-onset prediction. (b) Observation window up to 38 weeks with a 2-week prediction window (38–40 weeks), representing late-onset prediction. In these SHAP summary plots, each dot represents an individual pregnancy and colors indicate feature values (red = high, blue = low).

**Supplementary Figure 5.**
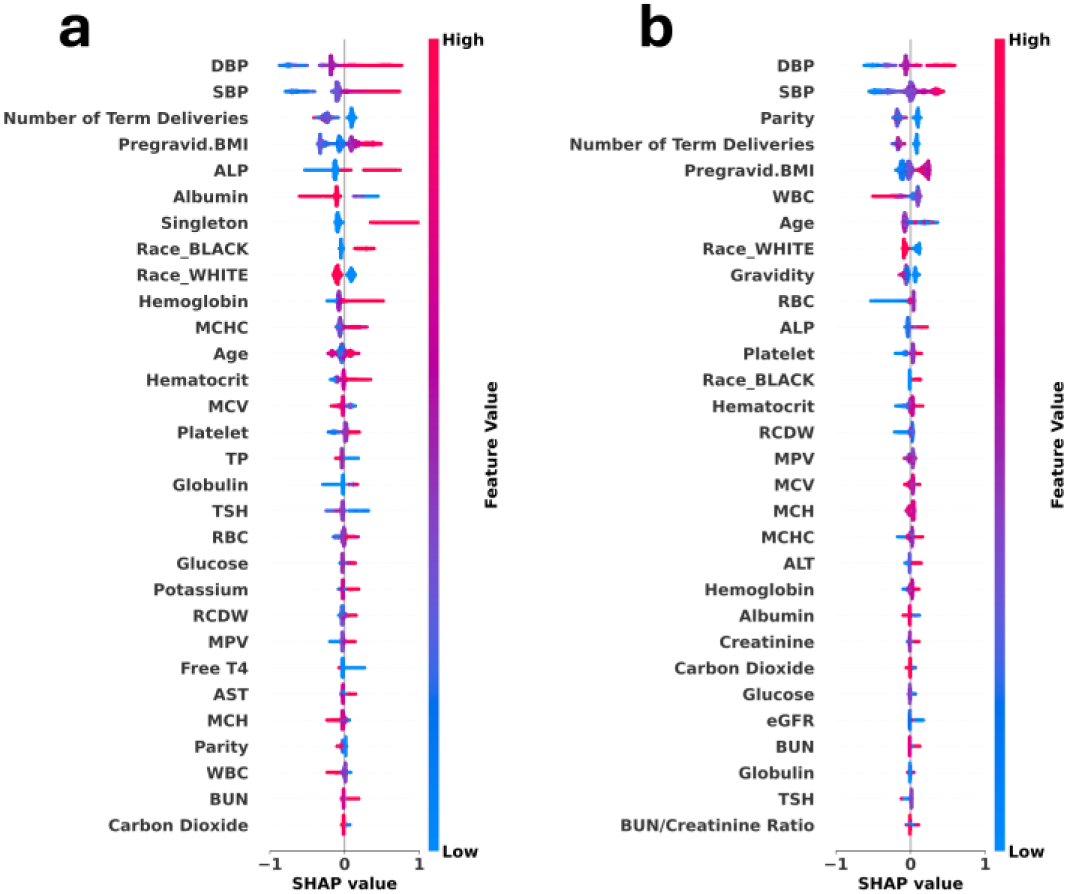
Top-30 global feature importance for PE prediction at early and late gestational stages. Global SHAP analyses were performed on XGBoost models (BMH direct transfer). (a) Observation window up to 32 weeks with a 2-week prediction window (32–34 weeks), representing early-onset prediction. (b) Observation window up to 38 weeks with a 2-week prediction window (38–40 weeks), representing late-onset prediction. In these SHAP summary plots, each dot represents an individual pregnancy and colors indicate feature values (red = high, blue = low).

**Supplementary Figure 6.**
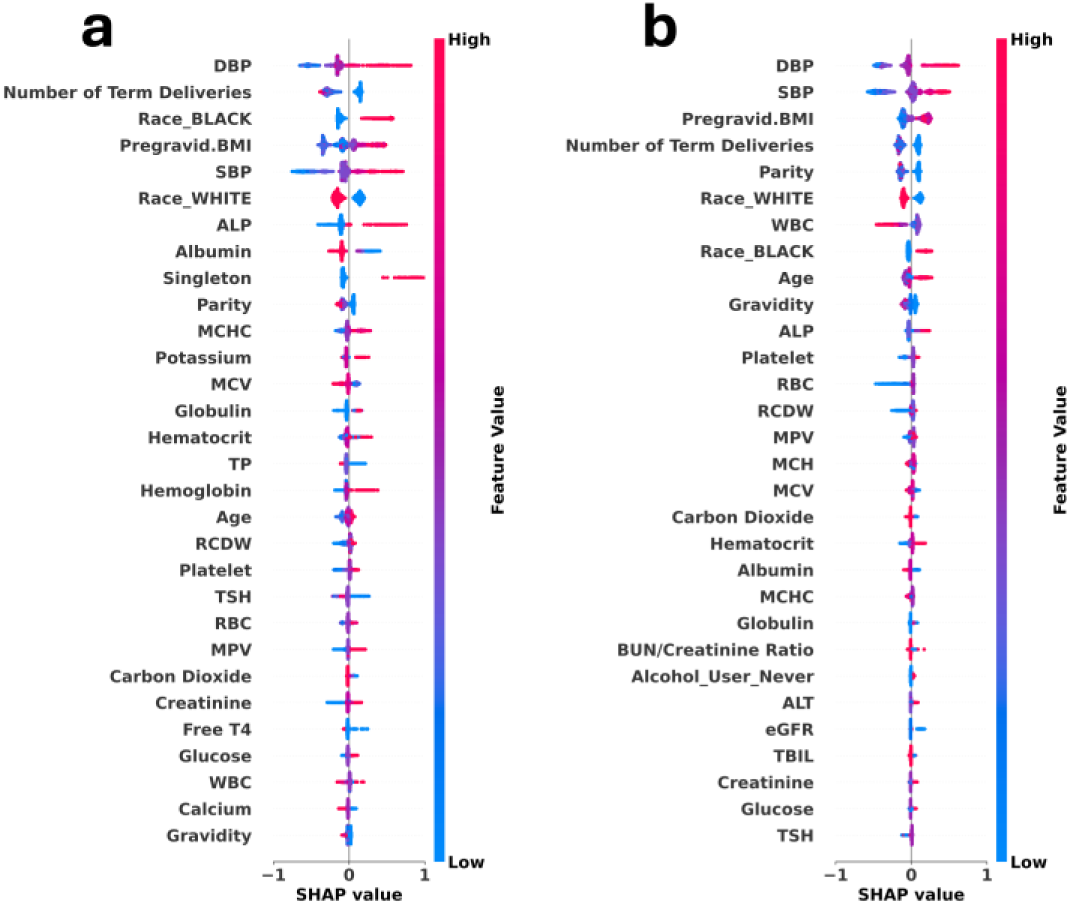
Top-30 global feature importance for PE prediction at early and late gestational stages. Global SHAP analyses were performed on XGBoost models (BMH fine-tuning). (a) Observation window up to 32 weeks with a 2-week prediction window (32–34 weeks), representing early-onset prediction. (b) Observation window up to 38 weeks with a 2-week prediction window (38–40 weeks), representing late-onset prediction. In these SHAP summary plots, each dot represents an individual pregnancy and colors indicate feature values (red = high, blue = low).

**Supplementary Figure 7.**
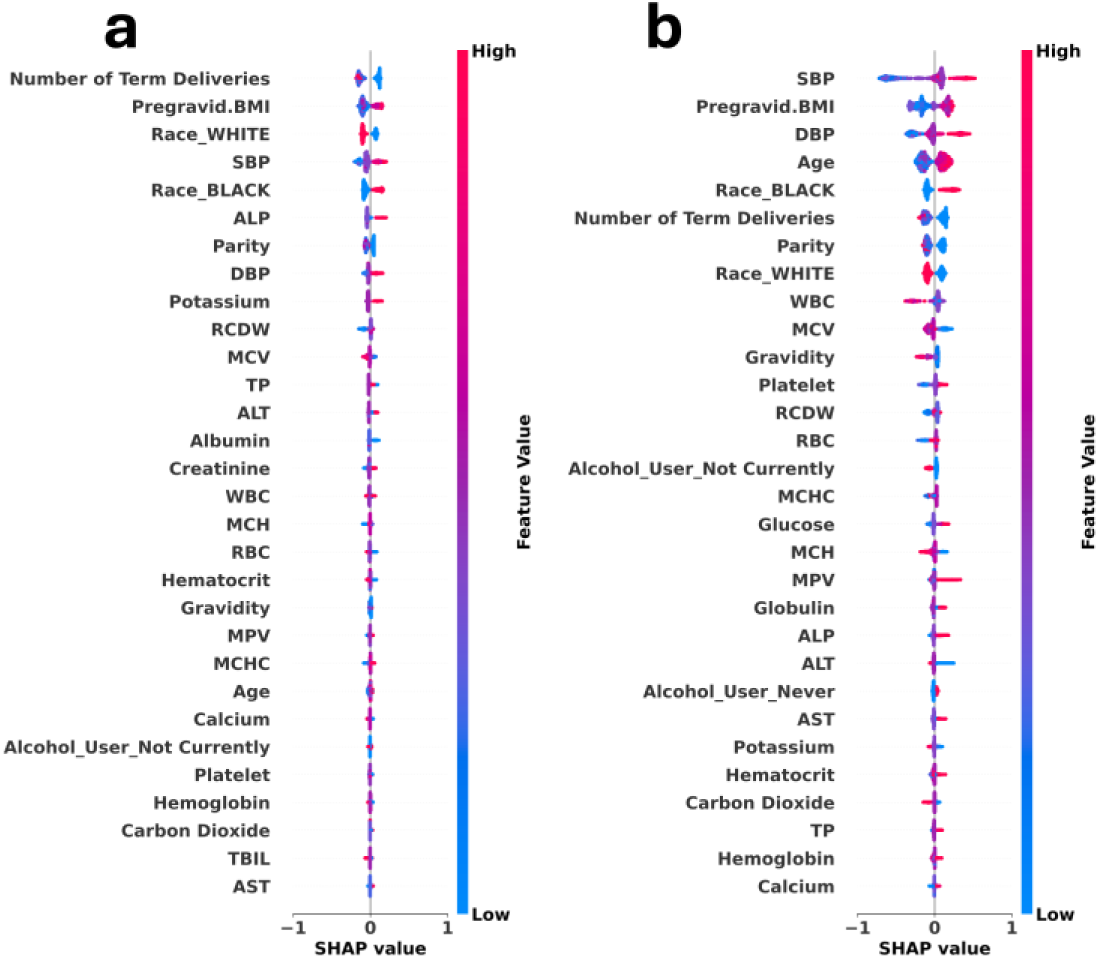
Top-30 global feature importance for PE prediction at early and late gestational stages. Global SHAP analyses were performed on XGBoost models (BMH retraining). (a) Observation window up to 32 weeks with a 2-week prediction window (32–34 weeks), representing early-onset prediction. (b) Observation window up to 38 weeks with a 2-week prediction window (38–40 weeks), representing late-onset prediction. In these SHAP summary plots, each dot represents an individual pregnancy and colors indicate feature values (red = high, blue = low).

